# Determinants of Neonatal Sepsis Admitted In Neonatal Intensive Care Unit At Public Hospitals Of Kaffa Zone, South West Ethiopia

**DOI:** 10.1101/2022.03.04.22271919

**Authors:** Melesse Belayneh, Gebremariam Getaneh, Alemaw Gebretsadik

## Abstract

**Background:** Neonatal sepsis is a systemic inflammatory response syndrome in the presence of infection during the first 28 days of life. Globally every year about 4 million children die in the first 4 weeks of life, of which 99% of the deaths occur in low and middle income countries and the most common causes of neonatal death in Ethiopia. Identification of the determinants for neonatal sepsis and treatment of newborns with sepsis is not adequate in low income countries like Ethiopia especially in southern part of the country.

**Objective:** To identify determinants of neonatal sepsis admitted in neonatal intensive care unit at public hospitals of Kaffa zone, south west Ethiopia 2021.

**Methods:** Institutional based unmatched case control study was conducted on a total sample of 248 (62 cases and 186 controls) in public Hospitals of kaffa zone from March to April 2021.The collected data were entered, coded and cleaned by Epidata version 3.1 and it was exported to SPSS version 25. Bi-variable and multivariable logistic regression was conducted. Variables with (p< 0.25) in bi-variable logistic regression analysis, were entered to multivariable logistic regression and then determinants which is statistical significant will be declared at P<0.05.

**Result:** A total of 248 (62 cases and 186 controls) were included in the study. variables like prolonged rupture of membrane ≥18 hours [AOR =5.13, 95%CI=1.38-19.05], meconium stained amniotic fluid[AOR =6.03, 95%CI=2.16-16.90], intra-partum fever [AOR =8.26, 95%CI=3.12-21.97], urinary tract infections [AOR=14,55, 95%CI=4.91-43.10], breast feeding after a hour [AOR =3.9, 95%CI=1.27-12.02], resuscitation [AOR =13.25, 95%CI=3.44-51.01], no chlorohexidine application [AOR =4.27, 95%CI=1.65-11.08] were significantly associated with neonatal sepsis.

**Conclusion and Recommendation:** Among the variables prolonged rupture of membranes, meconium stained amniotic fluid, intra-partum fever, UTI/STI, and not breast feeding with in a hour were maternal variables and resuscitation at birth and not application of chlorohexidine ointment on the umbilicus were neonatal variables that were found to be neonatal-related risk factors of neonatal sepsis. Infection prevention strategies and clinical management need to be strengthening and/or implementing by providing especial attention for the specified determinants.

## Introduction

Neonatal sepsis is a systemic inflammatory response syndrome in the presence of infection during the first 28 days of life(1, 2). It could be of bacterial, viral, fungal or rickettsia origin(2). It is estimated to cause twenty-six percent of all neonatal deaths worldwide(3).

Neonatal sepsis can be classified in to two subtypes depending upon whether the onset of symptoms is before 72 hours of life (early onset) or after 72 hours (late onset). Early onset neonatal sepsis (EONS) is caused by organisms prevalent in the maternal genital tract or in the delivery area(4, 5). Late onset neonatal sepsis (LONS) is caused by the organisms thriving in the external environment of the home or hospital. The infection is often transmitted through the hands of the care-providers(5, 6). It comprises abundant systemic infections of the neonate like septicemia, meningitis, pneumonia, arthritis, urinary tract infection but, it does not embrace muco-cutaneous infections like conjunctivitis and oral thrush. Neonatal infections in health care facilities are still a reason for inflated death and illness in Neonatal Intensive Care Unit (NICU) (5).

World health organization (WHO) has recognized seven clinical indicators [difficulty feeding, convulsions, movement only while stimulated /lethargy, respiratory rate of >60 breaths in a minute, chest in drawing, or axillary temperature of >37.5 °C or <35.5 °C and respiratory distress(7). Another study has additionally incorporated cyanosis and grunting(8).

Risk factors for EONS include prematurity, low birth weight, premature and prolonged rupture of membranes, maternal fever, uroinfection and chorioamnionitis whereas for LONS are invasive procedures such as resuscitation in delivery room, intubation, mechanical ventilation, central venous catheters, surgical procedures (5). As shown in different literatures, neonatal sepsis is caused by factors related to both maternal and neonatal factors (9-11). Delivery by caesarian section, male sex and prematurity have been showed as determinants of neonatal sepsis(11, 12). Maternal factors such as prolonged rupture of membrane, urinary tract infection, intra-partum fever, instrumental delivery and place delivery are significant predictors of neonatal sepsis (10, 13).

In other studies, use of endotracheal intubation, resuscitation at birth, surgery are significant determinants (11). Neonates born from women with meconium stained amniotic fluid, more than three times digital per vaginal examination and never attend antenatal care (ANC) are at higher risk for neonatal sepsis(14-16).

Neonatal sepsis is the commonest cause of neonatal morbidity and mortality. It is responsible for about thirty up to fifty percent of total neonatal deaths in developing countries(10). Globally every year about 4 million children die in the first 4 weeks of life, of which 99% of the deaths occur in low- and middle-income countries and of which 75% are considered as avoidable. Ethiopia continuous to struggle with a prevalence of about 42% or 81,000 newborn deaths every year(17).

Although several novel diagnostic markers have been discovered to detect onset of disease, at this moment, there is no proper diagnostic tool developed that can accurately diagnose sepsis. Promising results are emerging from recently identified cytokines, chemokines, cell surface antigens, physiomarkers, and metabolomics(18). The gold standard for establishing a diagnosis of neonatal sepsis is through culture. However, several factors

,including the small blood volumes obtained from neonates as well as maternal intra-partum antimicrobial exposure, can make the confirmation of sepsis in a neonate a diagnostic challenge(19)

Although recent medical advances have improved neonatal care many challenges remain in the diagnosis and management of neonatal infections. The diagnosis of neonatal sepsis is complicated by the frequent presence of non-infectious conditions that resemble sepsis, especially in preterm infants, and by the absence of optimal diagnostic tests. Efforts to reduce the rates of infection in this vulnerable population are one of the most important interventions in neonatal care(20).

Even though the presence of few studies regarding risk factors and etiology of neonatal sepsis, there are some contradicting or inconsistent findings on some predictors for neonatal sepsis, like prematurity, low birth weight and residence (10, 27). A study in Mexico showed, prematurity, low birth weight and residence as significant determinant of neonatal sepsis(11). On the contrary a study in Ethiopia showed that, all the three variables were not determinants of neonatal sepsis(10). Also a study in Wolayta Sodo showed that ANC visit, maternal age, maternal income as independent determinants of neonatal sepsis but in a study conducted in Jinka the above variables are not determinants(32, 33).

As far as literature searching showed, there is no study conducted in the study area regarding determinants of neonatal sepsis. Therefore, the aim of this study is to identify the determinants of neonatal sepsis among neonate admitted to the Neonatal Intensive Care Unit (NICU) in public hospitals of Kaffa zone, Southwest Ethiopia, 2021.

### Objective

To identify determinants of neonatal sepsis admitted in neonatal intensive care unit at public hospitals of Kaffa zone, south west Ethiopia 2021

## Methods and Materials

### Study design

Institutional based unmatched case control study design was conducted to assess the determinants of neonatal sepsis among neonates admitted at neonatal intensive care unit in public hospitals of Kaffa zone south west region of Ethiopia.

### Study Area and Period

The study was conducted from March 01 to April 10/2021 in Kafa zone south west region of Ethiopia. Kafa zone is one among 15 zones and 4 special woredas in SNNPRS with an elevation of 1450-2700 meters above sea level. The capital of kaffa is Bonga town which is located 450Km south west of Addis Ababa and 559 Km from regional city Hawassa. Kaffa zone is divided in to 12 woredas and 2 administrative towns. It has 1 General hospital which is Gebretsadik shawo General Hospital serving for more than 1,200,000 populations and 2 primary hospital (Wacha Primary Hospital and Telo Primary Hospital) serving for 211,000 and 84,000 populations respectively. According to information obtained from administrative offices of these hospitals, they provide different services in outpatient department, inpatient department and operation room theatre department. GSGH had 1 GP and 5 neonatal nurses with 10 functional beds while CPH had three nurses while TPH has two clinical nurses with three beds each in NICU with total annual neonatal admission of more than 2500 of which more than 850 was by neonatal sepsis in all hospitals of the catchment area. As a report from kaffa zone health department indicates the monthly neonatal sepsis new admission in March 2020 was 32 in GSGH, 13 in WPH, and 10 in TPH(55).

According to the 2020/21 annual report of Zonal health department, currently the population is 1,224,684 (male: 600,095 and female: 624,589 and from this 42,374 and 39,067 are live births and surviving infants respectively. The zone has up and down topography which is not suitable to get their health needs of the community. It is bordered by Jima zone in East and North, Konta special woreda and Basketo woreda in South, Benchimaji and Sheka zone in west.

### Population

#### Source population

All neonates admitted in neonatal intensive care unit in public hospitals of kaffa zone

#### Study population

All neonates admitted in neonatal intensive care unit in public hospitals of kaffa zone during the study period

#### Sample population

All randomly selected neonates for controls and consecutively selected neonates for cases in the neonatal intensive care unit.

### Variables

#### Dependent variable

Neonatal sepsis.

#### Independent Variables

Socio-demographic characteristics of neonates and parents (Age of mother, Ethnicity, Religion, Residence, Marital status, Educational status of mother, Occupation of mother, Neonatal age, Neonatal sex, Monthly income and Family size), maternal factors (Gravidity, Parity, ANC visit, Frequency of ANC, Place of delivery, Mode of delivery. Attendant of delivery, PROM, MSAF, Number of PV exam, Intra-partum fever, Foul smelling vaginal discharge, PIH /eclapsia, APH, UTI or STI and Breast feeding) and neonatal factors (Gestational age, Apgar score at 1st minute, Apgar score at 5th minute, Birth weight, Immediate cry, Birth asphyxia, Resuscitation, Birth trauma, Cord cut, Chlorohexidine application, Congenital anomalies and Invasive procedure) were the hypothesized determinants

### Inclusion and Exclusion criteria

#### Inclusion criteria

Neonates whose age less than one month(at the time of admission) with their mothers and admitted to neonatal intensive care unit of public hospitals of kaffa zone and fulfills the neonatal sepsis criteria for cases and neonates who admitted in NICU other than sepsis for controls.

#### Exclusion criteria

Neonates whose mothers had hearing impairments or unable to talk, abandoned neonates who were admitted in the hospital, and neonates whose cards had incomplete information, neonates whose mother is died, was excluded from the study in both controls and cases.

### Sample size estimation

The sample size was determined by using double population proportion formula in STATCALC program of the EPI INFO version 7.4.2 statistical software by considering double population proportion formula Proportion of maternal UTI/STI among controls is 14.9% and proportion of UTI/STI among cases is 33.8%; then by taking 95% CI, power of the study is 80%, control to case ratio of 3:1 the sample size becomes 227. After adding 10% non-respondent rate the final required sample will be 248 (62 cases and 186 controls) participants. Cases were neonates with sepsis and controls were neonates without sepsis who were admitted to NICU of the thee hospitals during the study period.

### Sampling procedure

Cases (diagnosed with sepsis) were selected through consecutive sampling technique from the neonates who were admitted in NICU until the required sample size achieved. The next immediate three corresponding controls were selected by simple random sampling (lottery method) at the same day and in the same neonatal intensive care unit. Proportional allocation of the sample was applied for the hospitals based on their case load (36 cases and 108 controls from GSGH, 15 cases and 45 controls from WPH, 11 cases and 33 controls from TPH).

#### Operational and term definitions

##### Case (Neonatal sepsis)

The established Integrated Management of Neonatal and Childhood Illness (IMNCI) clinical features, includes the presence of two or more of persistent fever (≥37.5 °C) or persistent hypothermia (≤35.5 °C) for more than one hour, fast breathing (≥60 breath per minute), severe chest in drawing, grunting, not feeding well, movement only when stimulated, bulged fontanel, convulsion, lethargic or unconsciousness along with ≥2 of the hematological criteria such as total leukocyte count (< 4000 or > 12,000 cells/mm3), absolute neutrophil count (< 1500 cells/mm3 or > 7500 cells/mm3), platelet count (< 150 or > 450 cells/mm3), and random blood sugar (< 40 mg/dl or > 125 mg/dl) will be used to diagnose neonatal sepsis cases(10).

##### Control

neonates with the diagnosis of non-sepsis case with their index mother(10).

##### Meconium stained amniotic fluid (MSAF)

will be considered if the amniotic fluid was green /brown in color or mixed with meconium, or appears meconium stained on the baby(54).

##### Prolonged rupture of membrane (PROM)

the time from membranes’ rupture to delivery more than 18 hours(54).

##### Invasive procedures

Procedures which have either of the following like Surgical procedures, Umbilical catheterization, Urinary catheterization, Nasogastric/oropharynx tube insertion, Endotracheal tube)(32).

### Data Collection materials and procedure

The questionnaire and checklist were developed after reviewing different literature which was related to determinants of neonatal sepsis. Then the designed questionnaires was changed from English to local language(kafinoono) and back to English to check the consistency of the questionnaire.. Training for supervisors and data collectors about data collection procedure was given for two days. Data were collected through a face to face interview of the index mothers with pretested structured interviewer administered questionnaires and reviewing neonates’ medical records (laboratory results:-CBC, CRP, ESR,) using checklist by trained experienced health professionals. They were interviewed about their socio-demographic characteristics, maternal factors and neonatal factors in each NICU in two primary hospitals and one general hospital. The data were collected from March to April 10/2021 by 4 BSc nurses and 1 MPH supervisor.

### Data Processing and Analysis

The filled questionnaires were checked for completeness and entered to Epidata version 3.1 then exported to SPSS version 25 for farther analysis. Descriptive statistics like frequency tables and percentages were used to describe the study participants in relation to relevant variables. The goodness of the model was assessed whether the required assumptions for the application of multivariate logistic regression will be fulfilled and the model will adequately fits the data (Hosmer and Lemeshow test was assessed). Both bi-variable and multivariable logistic regression models were used to identify factors which determinate neonatal sepsis. In order to minimize factors in multivariable analysis only variables that had showed p-value<0.25 on the Bi-variable logistic analysis were entered in to the multivariable logistic regression models. Odds Ratio and their 95% confidence intervals were computed and variables with p-value less than 0.05 in multivariable analysis were considered as statistically significant. Backward stepwise technique method will be used.

### Data Quality Assurance

Data quality assurance was implemented in all stage of the study starting from questionnaire designing, training, data collection and data entry. Pretest was done at Bonga health center which had NICU by taking 5% of sample size (3 cases and 9 controls) before the study period and the consistency of the questionnaire was checked. The questionnaire was objective based, logically sequenced and pre tested. The data collector and supervisor had received training on the objective of the study. The collected data was checked by the principal investigator on daily basis for any incompleteness. Data entry was conducted by principal investigator and other skilled personnel to minimize error during data entry.

### Ethical Consideration

Ethical clearance was obtained from Institutional Review Board (IRB) of Bahir Dar University College of Medicine and health science. Letter of permission was obtained from kaffa zone health department and each hospital Chie excutive office. The confidentiality of information had maintained by excluding personal identifiers and interview privately. Data were collected after securing informed written consent from every respondent.

## Result

### Socio-demographic characteristics of the respondent

This study involved a total of 248 neonates with their index mother among which 62 had sepsis (cases) and 186 was controls with 100% response rate. The study was intended to assess determinants of neonatal sepsis among neonates admitted in neonatal intensive care unit in kaffa zone hospitals. The mean age of index mothers was 28.06 years (SD ±5.82) with minimum age of 18 and maximum age of 42. The mean age of neonates was 5.04 days (SD ±6.09) with the age range of 1-27 days. Most of the participants were kaffa by ethnicity 43(69.4%) of cases and 139(74.7%) controls and 42(67.7%) of cases and 119 (64%) of controls were orthodox Christian. Among participants most of them were rural residents 38 (61.3%) of cases and 120 (64.5%) of controls). 82.3% cases and 83.9% of controls were married while 11(17.7%) of cases and 30(16.1%) of controls were educated above college level. Concerning maternal occupation, 75.8% of cases and 73.7% of controls were house wives while 19.4% of cases and of controls were go vernment employee. More than half of cases 36(58.1%) and controls 95(51%) were male neonate. The mean monthly income of cases and controls were 1970(SD ±1628) and 2197 (SD ±1191) ETB respectively(Table 1).

**Table 1:** Socio-demographic characteristics of mothers and their neonates for determinants of neonatal sepsis admitted in Neonatal Intesnive Care unit of public hospitals in Kaffa zone southwest Ethiopia 2021.

| Variables | Categories | Case n =62(%) | Control =186(%) | Total 248(%) |
| --- | --- | --- | --- | --- |
| Age of mother | 15-20 | 8(12.9) | 14(7.5) | 22(8.9) |
|  | 21-34 | 44(71.0) | 139(74.1) | 183(73.8) |
| | ≥35 | 10(16.1) | 33(17.7) | 43(17.3) |
| Ethnicity | Kafa | 43(69.4) | 139(74.7) | 182(73.4) |
|  | Oromo | 6(9.7) | 33(17.7) | 39(15.7) |
|  | Amhara | 11(17.7) | 14(7.5) | 25(10.1) |
|  | Konta | 2(3.2) | 0(0.0) | 2(0.8) |
|  | others | 0(0.0) | 0(0.0) | 0(0.0) |
| Religion | Orthodox | 42(67.7) | 119(64.0) | 161(64.9) |
|  | Protestant | 16(28.8) | 55(29.6) | 71(28.6) |
|  | Muslim | 4(6.5) | 12(6.5) | 16(6.5) |
| Residence | Urban | 24(38.7) | 66(35.5) | 90(36.3) |
|  | Rural | 38(61.3) | 120(64.5) | 158(63.7) |
| Marital status | Single | 8(12.9) | 17(9.1) | 25(10.1) |
|  | Married | 51(82.3) | 156(83.9) | 207(83.5) |
|  | Divorced | 2(3.2) | 7(3.8) | 9(3.6) |
|  | Widowed | 1(1.6) | 6(3.2) | 7(2.8) |
| Educational status of mother | Illiterate | 31(50.0) | 94(50.5) | 125(50.4) |
|  | Read and write | 10(16.1) | 40(21.5) | 50(20.2) |
|  | Elementary school | 5(8.1) | 11(5.9) | 16(6.5) |
|  | Secondary school | 5(8.1) | 11(5.9) | 16(6.5) |
|  | College/above | 11(17.7) | 30(16.1) | 41(16.5) |
| Occupation of mother | House wife | 47(75.8) | 137(73.7) | 184(74.2) |
|  | Gov't employee | 12(19.4) | 36(19.4) | 48(19.4) |
|  | Private institution | 2(3.2) | 5(2.7) | 7(2.8) |
|  | Self-employee | 0(0.0) | 4(2.2) | 4(1.6) |
|  | Daily laborer | 1(1.6) | 4(2.2) | 5(2.0) |
| Neonatal age | 1-3 | 43(69.3) | 107(57.5) | 150(60.5) |
|  | >3 | 19(30.6) | 79(42.5) | 98(39.5) |
| Neonatal sex | Male | 36(58.1) | 95(51.1) | 131(52.8) |
|  | Female | 26(41.9) | 91(48.9) | 117(47.2) |
| Monthly income | <3500 | 49(79.0) | 147(79.0) | 196(79.0) |
|  | >=3500 | 13(21.0) | 39(21.0) | 52(21.0) |
| Family size | <5 | 39(62.9) | 107(57.5) | 146(58.9) |
|  | >=5 | 23(37.1) | 79(42.5) | 102(41.1) |

### Maternal factors

According to this study 5(8.1%) of cases and 21(11.3%) of controls were Primigravida while 67.7% of cases and 64.5% of controls were multipara index mothers. Nearly two-third (63%) of cases and 166(89.2%) of controls were visited health facility for ANC service among them who visited for ANC 66% of cases and 70.4% of controls were visited more than three times. Among the participants, 23 (37.1%) of cases and 40.9% of controls,30(40.4%) of cases and 43.5% of controls,9(14.5%) of cases and 15.6% of controls were delivered at hospital, health center and home respectively. Three fourth (75.8%) of cases and 87.6% of controls were delivered through SVD while 8.1% of cases and 4.8% of controls were CS delivery and most deliveries were attended by health professionals. Twenty two (35.5%) of cases and 7% of controls had rupture of membranes greater than or equal to 18 hours. Two third (65.6%) of cases and nearly one-fourth (24.2%) controls had amniotic fluid discoloration while nearly half 48.4% of cases and 22.6 % controls had foul smelling amniotic fluid. Thirty nine (62.9%) of cases and 139(74.7%) of controls had per-vaginal examination of three times or less. Among respondents 46(74.2%) of cases and 41(22%) of controls had history of fever during the index pregnancy. Twenty two (35.5%) of cases and 27.4% of cases had PIH/eclapsia and APH respectively. One-fourth (19.9%) of controls 74.2% of case had UTI/STI during their index pregnancy while 40(64.5%) of cases 165(88.7%) of controls start breast feeding within one hour(Table 2).

**Table 2:** Maternal factors for determinants of neonatal sepsis admitted in Neonatal Intensive Care Unit of public hospitals in Kaffa zone southwest Ethiopia 2021.

| Variables | Categories | Case n=62(%) | Control n=186(%) | Total n=248(%) |
| --- | --- | --- | --- | --- |
| Gravidity | Primigravida | 5(8.1) | 21(11.3) | 26(10.5) |
|  | Multigravida | 57(91.9) | 165(88.7) | 222(89.5) |
| Parity | Nulipara | 6(9.7) | 21(11.3) | 27(10.9) |
|  | Paraone | 14(22.6) | 45(24.2) | 59(23.8) |
|  | Multipara | 42(67.7) | 120(64.5) | 162(65.3) |
| ANC visit | Yes | 39(62.9) | 166(89.2) | 205(82.7) |
|  | No | 23(37.1) | 20(10.8) | 43(17.3) |
| Frequency of ANC | <=3 | 21(33.9) | 55(29.6) | 76(30.6) |
|  | >3 | 41(66.1) | 131(70.4) | 172(69.4) |
| Place of delivery | Home | 9(14.5) | 29(15.6) | 38(15.3) |
|  | Health center | 30(48.4) | 81(43.5) | 111(44.8) |
|  | Hospital | 23(37.1) | 76(40.9) | 99(39.9) |
| Mode of delivery | SVD | 47(75.8) | 163(87.6) | 210(84.7) |
|  | Instrumental | 10(16.1) | 14(7.5) | 24(9.7) |
|  | CS | 5(8.1) | 9(4.8) | 14(5.6) |
| Attendant of delivery | TBA | 7(11.3) | 28(15.1) | 35(14.1) |
|  | HEW | 0(0.0) | 0(0.0) | 0(0.0) |
|  | Health professional | 55(88.7) | 158(84.9) | 213(85.9) |
| PROM | <12 | 38(61.3) | 171(91.9) | 209(84.3) |
|  | 12-18 | 2(3.2) | 2(1.1) | 4(1.6) |
|  | >=18 | 22(35.5) | 13(7.0) | 35(14.1) |
| MSAF | Yes | 40(65.6) | 45(24.2) | 85(34.3) |
|  | No | 21(34.4) | 141(75.8) | 162(65.3) |
| Number of PV exam | <=3 | 39(62.9) | 139(74.7) | 178(71.8) |
|  | >3 | 23(37.1) | 47(25.3) | 70(28.2) |
| Intra-partum fever | Yes | 46(74.2) | 41(22.0) | 87(35.1) |
|  | No | 16(25.8) | 145(78.0) | 161(64.9) |
| Foul smelling vaginal discharge | Yes | 30(48.4) | 42(22.6) | 72(29.0) |
|  | No | 32(51.6) | 144(77.4) | 176(71.0) |
| PIH /eclapsia | Yes | 22(35.5) | 25(13.4) | 47(19.0) |
|  | No | 40(64.5) | 161(86.6) | 201(81.0) |
| APH | Yes | 17(27.4) | 41(22.0) | 58(23.4) |
|  | No | 45(72.6) | 145(78.0) | 190(76.6) |
| UTI or STI | Yes | 46(74.2) | 37(19.9) | 83(33.5) |
|  | No | 16(25.8) | 149(80.1) | 165(66.5) |
| Breast feeding | <60 minute | 40(64.5) | 165(88.7) | 205(82.7) |
|  | >60 minute | 22(35.5) | 21(11.3) | 43(17.3) |

APH ante partum hemorrhage, PPH postpartum hemorrhage, GDM gestational diabetes mellitus, PIH pregnancy induced hypertension, PROM premature rupture of membrane and UTI Urinary Tract Infection

### Neonatal factors

According to this study 35(56.5%) of cases and 115(61.8%) of controls were in between 37-42 completed weeks of gestational age. The 1^st^ minute and 5^th^ minute apgar score <7 among case neonates were 24.2% and 9.2% respectively while the 1^st^ and 5^th^ minute apgar score <7 in controls were 9.7% and 1.6% respectively. More than 90% of cases and controls were in the normal range of birth weight. Forty one 66.1% of cases and 166(89.2%) of controls cries immediately after birth where as 34% of cases and 9.7% of controls developed birth asphyxia. One third of cases and 8.1% of controls were resuscitated after birth and more than 92% of case and controls had cord cut with new blade. Twenty two (35.5%) of cases 134(72%) of controls had history of chlorohexidine application for neonates where as 4.8% and 6% cases and controls had had invasive procedure(Table 3).

**Table 3.** Neonatal factors for the study of determinants of neonatal sepsis admitted in Neonatal Intesnive Care Unit of public hospitals in Kaffa zone southwest Ethiopia 2021.

| Variables | Categories | Case n=62(%) | Control n=186(%) | Total n=248(%) |
| --- | --- | --- | --- | --- |
| Gestational age | <37 | 27(43.5) | 71(38.2) | 98(39.5) |
|  | 37-42 | 35(56.5) | 115(61.8) | 150(60.5) |
|  | >42 | 0(0.0) | 0(0.0) | 0(0.0) |
| Apgar score at 1<sup>st</sup> minute | <7 | 15(24.2) | 18(9.7) | 33(13.3) |
|  | ≥7 | 47(75.8) | 168(90.3) | 215(86.7) |
| Apgar score at 5<sup>th</sup> minute | <7 | 6(9.7) | 3(1.6) | 9(3.6) |
|  | ≥7 | 56(90.3) | 183(98.4) | 239(96.4) |
| Birth weight | <2500 | 6(9.7) | 11(5.9) | 17(6.9) |
|  | ≥2500 | 56(90.3) | 175(94.1) | 231(93.1) |
| Immediate cry | Yes | 41(66.1) | 166(89.2) | 207(83.5) |
|  | No | 21(33.9) | 20(10.8) | 41(16.5) |
| Birth asphyxia | Yes | 21(33.9) | 18(9.7) | 39(15.7) |
|  | No | 41(66.1) | 168(90.3) | 209(84.3) |
| Resuscitation | Yes | 21(33.9) | 15(8.1) | 36(14.5) |
|  | No | 41(66.1) | 171(91.9) | 212(85.5) |
| Birth trauma | Yes | 4(6.5) | 8(4.3) | 12(4.8) |
|  | No | 58(93.5) | 178(95.7) | 236(95.2) |
| Cord cut | New blade | 57(91.9) | 173(93.0) | 230(92.7) |
|  | Old blade | 5(8.1) | 13(7.0) | 18(7.3) |
| Chlorohexidine application | Yes | 22(35.5) | 134(72.0) | 156(62.9) |
|  | No | 40(64.5) | 52(28.0) | 92(37.1) |
| Congenital anomalies | Yes | 2(3.2) | 13(7.0) | 15(6.0) |
|  | No | 60(96.8) | 173(93.0) | 233(94.0) |
| Invasive procedure | Yes | 3(4.8) | 11(5.9) | 14(5.6) |
|  | No | 59(95.2) | 175(94.1) | 234(94.4) |

### Health facility related factors

All neonates admitted in which all hospitals implemented infection prevention practice in the neonatal intensive care unit while 82.3% of case and control neonates were admitted in hospitals which had trained professional on infection prevention and control practice. Thirty nine (62.9%) of cases and 102(54.8%) of controls lives with in 5km or less and their common means of transportation for rural residents were hoarse(Table 4).

**Table 4:** Health facility related factors for the study of determinants of neonatal sepsis admitted in Neonatal Intensive Care Unit of public hospitals in Kaffa zone southwest Ethiopia 2021.

| Variables | Categories | Case n=62(%) | Control n= 186(%) | Total n=248(%) |
| --- | --- | --- | --- | --- |
| Infection prevention implemented | Yes | 62(100.0) | 186(100.0) | 248(100.0) |
|  | No | 0(0.0) | 0(0.0) | 0(0.0) |
| Trained professional on IPC | Yes | 51(82.3) | 153(82.3) | 204(82.3) |
|  | No | 11(17.7) | 33(17.7) | 44(17.7) |
| Distance of HF | ≤5 | 39(62.9) | 102(54.8) | 141(56.9) |
|  | >5 | 23(37.1) | 84(45.2) | 107(43.1) |
| Means of transport | Car | 32(51.6) | 85(45.7) | 117(47.2) |
|  | Hoarse | 25(40.3) | 85(45.7) | 110(44.4) |
|  | Foot | 5(8.1) | 16(8.6) | 21(8.5) |

### Determinants of Neonatal Sepsis Admitted In Neonatal Intensive Care

In bi-variable logistic regression analysis result ANC visit, mode of delivery, duration of PROM, meconium stained amniotic fluid, frequency of PV exam, intra-partum fever, foul smelling amniotic fluid, PIH, UTI, breast feeding, gestational age, apgar score <7 at 1^st^ and 5^th^ minute, immediate cry, asphyxia, resuscitation, chlorohexidine application were showed association with neonatal sepsis(P<0.25). then applying multivariable logistic regression maternal PROM ≥18 hours, MSAF, intra-partum fever, history of UTI/STI, breast feeding after 60 minute, neonatal resuscitation at birth and chlorohexidine application was significantly associated with neonatal sepsis.

In multivariable logistic regression analysis, neonates born from women who had prolonged rupture of membrane ≥18 hours had five times more likely to develop neonatal sepsis than that of women who had PROM <12 hours [AOR =5.13; 95% CI(1.38, 19.05)]. Those neonates born from mothers who had MSAF were six times more likely to suffer from neonatal sepsis than their counter parts [AOR =6.03; 95%CI(2.16, 16.90)]. Intra-partum fever was also had a significant association with neonatal sepsis, mothers who had history of intra-partum fever were eight times more risk for their neonates to develop neonatal sepsis than mothers who had no fever [AOR =8.26; 95%CI(3.12, 21.97)].

In this study neonatal sepsis was strongly associated with maternal history of UTI/STI, those neonates born from mothers who had history of UTI/STI were fourteen times more likely to have neonatal sepsis than their counter parts [AOR =14,55; 95%CI(4.91, 43.10)]. Mothers who breast feed their neonate after 1 hour were nearly four times more likely to suffer from neonatal sepsis than that of neonates who breast feed within 1 hour [AOR =3.9; 95%CI(1.27, 12.02)]. Resuscitation of the newborn was also had significant association with neonatal sepsis, resuscitated neonates were thirteen times more likely to have neonatal sepsis than their counter parts [AOR =13.25; 95%CI(3.44, 51.01)]. Those neonates who were not got chlorohexidine ointment on their umbilicus had four times more likely to develop neonatal sepsis than neonates who had got chlorohexidine application [AOR =4.27; 95%CI(1.65, 11.08)] (Table 5).

**Table 5:**
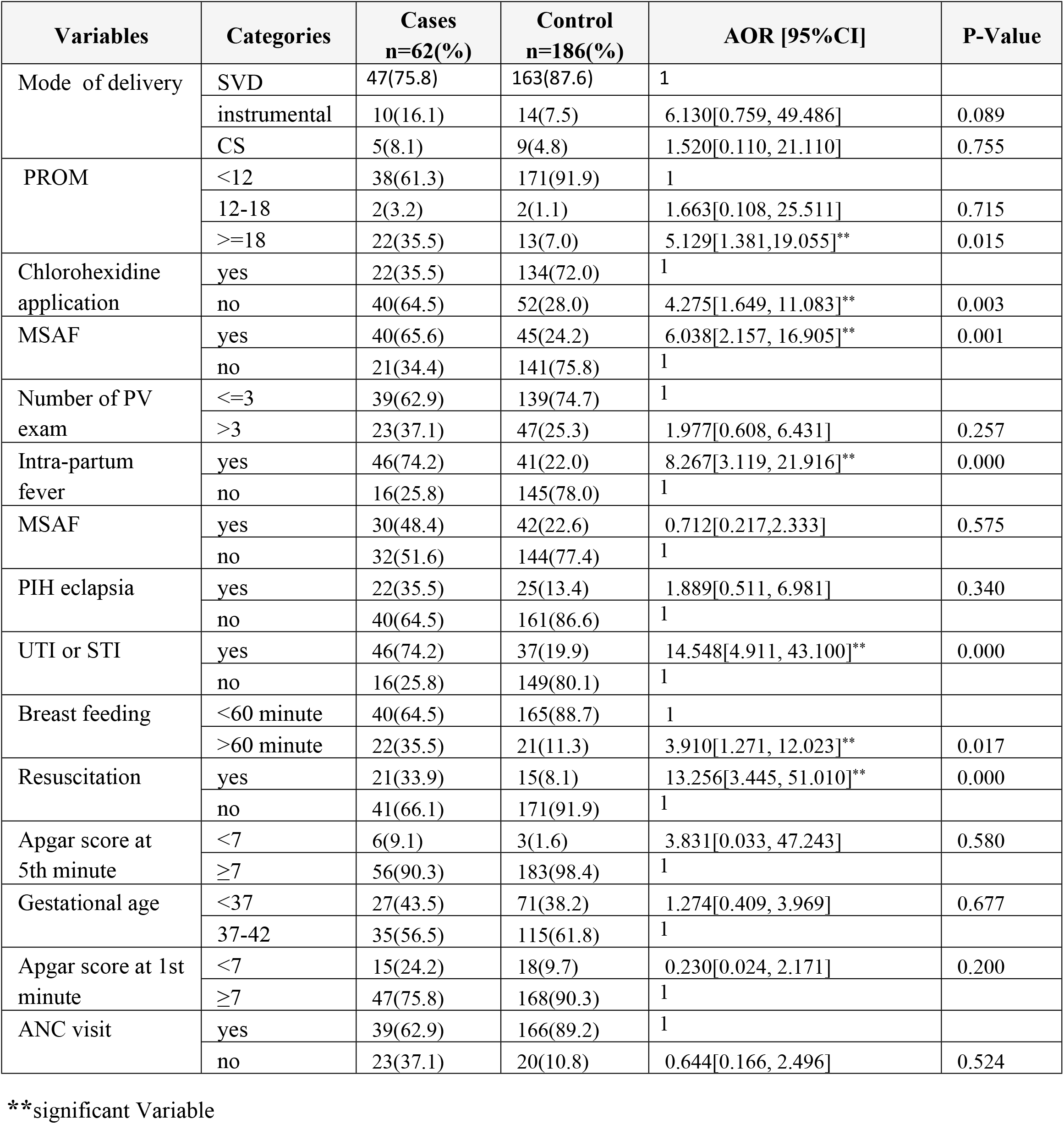
multivariable logistic regression analysis result determinants of neonatal sepsis admitted in Neonatal Intensive Care Unit of public hospitals in Kaffa zone southwest Ethiopia 2021.

| Variables | Categories | Cases<br>n=62(%) | Control<br>n=186(%) | AOR [95%CI] | P-Value |
| --- | --- | --- | --- | --- | --- |
| Mode of delivery | SVD | 47(75.8) | 163(87.6) | 1 |  |
|  | instrumental | 10(16.1) | 14(7.5) | 6.130[0.759, 49.486] | 0.089 |
|  | CS | 5(8.1) | 9(4.8) | 1.520[0.110, 21.110] | 0.755 |
| PROM | <12 | 38(61.3) | 171(91.9) | 1 |  |
|  | 12-18 | 2(3.2) | 2(1.1) | 1.663[0.108, 25.511] | 0.715 |
|  | >=18 | 22(35.5) | 13(7.0) | 5.129[1.381, 19.055]** | 0.015 |
| Chlorohexidine application | yes | 22(35.5) | 134(72.0) | 1 |  |
|  | no | 40(64.5) | 52(28.0) | 4.275[1.649, 11.083]** | 0.003 |
| MSAF | yes | 40(65.6) | 45(24.2) | 6.038[2.157, 16.905]** | 0.001 |
|  | no | 21(34.4) | 141(75.8) | 1 |  |
| Number of PV exam | <=3 | 39(62.9) | 139(74.7) | 1 |  |
|  | >3 | 23(37.1) | 47(25.3) | 1.977[0.608, 6.431] | 0.257 |
| Intra-partum fever | yes | 46(74.2) | 41(22.0) | 8.267[3.119, 21.916]** | 0.000 |
|  | no | 16(25.8) | 145(78.0) | 1 |  |
| MSAF | yes | 30(48.4) | 42(22.6) | 0.712[0.217, 2.333] | 0.575 |
|  | no | 32(51.6) | 144(77.4) | 1 |  |
| PIH eclapsia | yes | 22(35.5) | 25(13.4) | 1.889[0.511, 6.981] | 0.340 |
|  | no | 40(64.5) | 161(86.6) | 1 |  |
| UTI or STI | yes | 46(74.2) | 37(19.9) | 14.548[4.911, 43.100]** | 0.000 |
|  | no | 16(25.8) | 149(80.1) | 1 |  |
| Breast feeding | <60 minute | 40(64.5) | 165(88.7) | 1 |  |
|  | >60 minute | 22(35.5) | 21(11.3) | 3.910[1.271, 12.023]** | 0.017 |
| Resuscitation | yes | 21(33.9) | 15(8.1) | 13.256[3.445, 51.010]** | 0.000 |
|  | no | 41(66.1) | 171(91.9) | 1 |  |
| Apgar score at 5th minute | <7 | 6(9.1) | 3(1.6) | 3.831[0.033, 47.243] | 0.580 |
|  | ≥7 | 56(90.3) | 183(98.4) | 1 |  |
| Gestational age | <37 | 27(43.5) | 71(38.2) | 1.274[0.409, 3.969] | 0.677 |
|  | 37-42 | 35(56.5) | 115(61.8) | 1 |  |
| Apgar score at 1st minute | <7 | 15(24.2) | 18(9.7) | 0.230[0.024, 2.171] | 0.200 |
|  | ≥7 | 47(75.8) | 168(90.3) | 1 |  |
| ANC visit | yes | 39(62.9) | 166(89.2) | 1 |  |
|  | no | 23(37.1) | 20(10.8) | 0.644[0.166, 2.496] | 0.524 |

**\*\*significant Variable**

## Discussion

The finding of this study showed that the duration of rupture of membrane ≥18 hours, meconium stained amniotic fluid, intra-partum maternal fever, history of UTI/STI, and breast feeding after 1 hour were independent maternal factors for neonatal sepsis whereas neonatal resuscitation and chlorohexidine application were identified as neonatal determinants of neonatal sepsis.

In this study prolonged rupture of membrane ≥18 hours was significantly associated with neonatal sepsis, neonates born from women who had prolonged rupture of membrane ≥18 hours had five times more likely to develop neonatal sepsis than that of women who had PROM <12 hours. This finding is in line with studies conducted in Mekele, Mexico, Jinka, Nepal, USA, China, Pakistan, Northwest Ethiopia (10, 11, 33, 35, 47, 48, 49, 53). These may be due to birth canal is colonized with aerobic and anaerobic pathogens that might cause in ascending amniotic fluid infection and colonization of the neonate at birth. Early rupture of membrane increases the chance of ascending microorganisms from the birth canal into the amniotic sac causing chorioamnionitis and fetal compromise which frequently leads the new born to infection in utero. Mother to fetus transmission of bacterial agents that infect the amniotic fluid and birth canal may occur in uterus more commonly during labor and delivery which results in neonatal sepsis.

Meconium stained amniotic fluid had also a significant association with neonatal sepsis. Those neonates born from mothers who had MSAF were six times more likely to suffer from neonatal sepsis than neonates who had no MSAF. These finding is comparable with a study conducted in Mexico, south Africa, Uganda, Nepal, Ghana (11, 15, 16, 35, 45). This may be due to that, neonates delivered from women with meconium stained amniotic fluid are more liable to aspirate it and fill smaller air ways and alveoli in the lung. And it increases the multiplication of microbes that cause sepsis.

Intra-partum fever was also had a significant association with neonatal sepsis, mothers who had history of intra-partum fever were eight times more risk for their neonates to develop neonatal sepsis than mothers who had no fever. These finding is consistent with studies conducted in Mekele, Wolayta sodo, and Pakistan (32). This might be because of the reason that fever is a sign of local or systemic infections such as chorioamnionitis and urinary tract infection, which are frequently transmitted to the baby in the uterus or during passage through the canal This results in hematogenious spread and vertical transmission of pathogens to the newborn before or during labor and delivery which further results in neonatal sepsis.

In this study neonatal sepsis was strongly associated with maternal history of UTI/ STI, those neonates born from mothers who had history of UTI/STI were fourteen times more likely to have neonatal sepsis than mothers who had no UTI/STI. This is in line with Mekele, Uganda, Jinka, central Ethiopia, and Ghana which showed UTI was significant determinants of neonatal sepsis (10, 16, 33, 34, 45). This might be due to, the wall of birth canal in women with urinary tract infection are colonized with pathogens. The most common microorganisms that cause neonatal sepsis are found across the birth canal and possibly increase the risk while the newborn was born and pass through the vaginal wall.

Breast feeding was also had association with neonatal sepsis, Mothers who breast feed their neonate after 1 hour were nearly four times more likely to suffer from neonatal sepsis than that of neonates who breast feed within 1 hour. These finding is consistent with a study conducted in Wolayta sodo (32). Early initiation of breast feeding has different health benefit like increase ability to defense infection and increase the survival rate of children because colostrum is the first milk that is very important for newborns in protecting infection since rich in immunoglobin G that has great role in disease resistance.

Resuscitation of the newborn had significant association with neonatal sepsis; resuscitated neonates were thirteen times more likely to have neonatal sepsis than their counter parts. This finding is comparable with studies done in Tanzania, Ghana, NW Ethiopia, and another Northwest Ethiopia (3, 45, 53, 54). These findings might be due to the fact that, if procedure of resuscitation is done forcefully, it may cause laceration to the susceptible and easily breakable mucous membrane of the neonate and serve as a route of entry for pathogens from unsterile equipment. It may also lead microbes into the lower air way of the newborn with an immature immune system. This is due to the lumen of airways of the neonate is too narrow, and respiratory secretions are copious compared to older children who could predispose to easily destruction of smaller air sacs and sepsis. Moreover, resuscitation might be done with unsterile equipment, which could introduce microbes into the lungs of the neonate whose immune system is not yet well developed.

Chlorohexidine application had also significant association with the occurrence of neonatal sepsis. Neonates who had no application of chlorohexidine were having a significant risk for neonatal sepsis. This may be due to that umbilical cord is the main portal of entry for microorganisms that cause neonatal sepsis(56, 57).

## Limitation of the study

Since the study was done on admitted neonates, thus results might lack generalizability to the entire population.

## Conclusion

Among the variables prolonged rupture of membranes, meconium stained amniotic fluid, intra-partum fever, UTI/STI, and not breast feeding with in a hour were maternal variables and resuscitation at birth and not application of chlorohexidine ointment on the umbilicus were neonatal variables that were found to be neonatal-related risk factors of neonatal sepsis. Infection prevention strategies and clinical management need to be strengthening and/or implementing by providing especial attention for the specified determinants.

## Recommendation

Health professionals who are working in NICU and delivery room should use aseptic technique, clean sterilize or disinfect instruments and equipment, and routinely clean the neonatal intensive care unit. Special attention and follow up should be given for neonates who born from mothers who had history of PROM, MSAF, UTI/STI and intra-partum fever. Screen mothers for UTI and STI and treat accordingly.Mobilize and support early health-seeking behavior for health problems during pregnancy, like seeking care for UTI/STI. Encourage mothers to breastfeed their newborns and initiate as early as possible. Give regular infection prevention, health education and promotion on maternal and neonatal health.

## Data Availability

Melesse B. conceptualized, data collection, analyze and interpret the data. the manuscript prepared.

## Availability of data and materials

All the data are available in the manuscript.

## Abbreviations

ANC: Antenatal Care
AOR: Adjusted Odds Ratio
APGAR: Activity, Pulse, Grimace, Appearance, Respiration
APH: Antepartum Hemorrhage
BW: Birth Weight
CBC: Complete Blood Count
CI: Confidence Interval
COR: Crude Odds Ratio
CRP: C-Reactive Protein
EDHS: Ethiopian Demographic and Health Survey
EONS: Early Onset Neonatal Sepsis; Integrated Management of Neonatal and Childhood Illness
GSGH: Gebretsadik Shawo General Hospital
LONS: Late onset neonatal sepsis
MSAF: Meconium Stained Amniotic Fluid
NGT: Nasogastric Tube
NMR: Neonatal Mortality Rate
NICU: Neonatal intensive care unit
PIH: Pregnancy Induced Hypertension
PROM: Premature Rupture of Membrane
PSBI: Possible Serious Sever Bacterial Infection
SDG: -Sustainable Development Goal
SPSS: Statistical package for Social Science
SSA: Sub-Saharan Africa
UTI: Urinary Tract Infection
TPH: Telo Primary Hospital
WHO: World Health Organization
WPH: Wacha Primary Hospital

## Acknowledgments

The authors acknowledged Bahir Dar University, Wacha Primary Hospital, Telo Primary Hospital, Gebretsadik Shawo General Hospital, and data collectors.

## Contributions

MB is the pioneer to draft the manuscript. GG & AG are research assistant who has been actively engaged in each step in the research process. MB, GG & AG were involved in the conception and design of the whole research. MB, GG & AG is involved in the analyzed and interpreted of findings. All authors have read and approved their ownership the manuscript.

## Ethics declarations

### Ethics approval and consent to participate

Ethical clearance was obtained from Bahir Dar University, College of Medicine and Health sciences, Institutional Ethics Review Board (IRB) with letter refrence number (IRB 2345/2021). Informed consent was taken from each participant.

### Consent for publication

Not applicable

